# Migraine and Early-onset Ischemic Stroke: A Mendelian Randomization Study

**DOI:** 10.1101/2024.08.01.24311334

**Authors:** Peng-Peng Niu, Rui Zhang, Shuo Li, Yu-Sheng Li

## Abstract

**Objective:** Through the utilization of the data specifically related to early-onset ischemic stroke, we aimed to investigate the causal effects of migraine and its subtypes on the risk of early-onset ischemic stroke using the two-sample Mendelian randomization method.

**Methods:** Genetic instrumental variables were acquired from two sources with the largest sample sizes available. Summary data for early-onset ischemic stroke was acquired from a study encompassing individuals aged 18 to 59 years, comprising 16,730 cases and 599,237 non-stroke controls. The random-effects inverse variance weighted method was used as the primary analysis approach.

**Results:** The Mendelian randomization analysis revealed no association between overall migraine and migraine without aura with the risk of early-onset ischemic stroke. However, migraine with aura showed a suggestive association with an elevated risk of early-onset ischemic stroke, with odds ratios of 1.114 (95% confidence interval = 1.005 to 1.236, p-value = 0.040) and 1.062 (95% confidence interval = 1.002 to 1.126, p-value = 0.042) based on instruments from two independent sources. The odds ratio was 1.074 (95% confidence interval = 1.022 to 1.130, p-value = 0.005) based on instruments from both two sources. No evidence of heterogeneity or horizontal pleiotropy was found. Furthermore, a positive genetic correlation was found between migraine with aura and early-onset ischemic stroke (genetic correlation = 0.208, 95% confidence interval = 0.038 to 0.377, p-value = 0.016). By contrast, migraine with aura was not related to ischemic stroke of all adults.

**Conclusion:** This study provides evidence of a causal relationship between migraine with aura and the risk of early-onset ischemic stroke.

## Introduction

Migraine, a prevalent neurological disorder, is characterized by recurrent and often severe headaches, frequently accompanied by a range of associated symptoms. It has a global age-standardized prevalence of approximately 14%.^1^ Observational studies have shown that there was an increased risk of ischemic stroke for individuals who are afflicted with migraine, especially for those with migraine with aura. However, the preceding Mendelian randomization (MR) studies have yet to succeed in confirming the causal connection between them.^2, 3^ The lack of conclusive MR findings may be due to migraine’s potential association with early-onset rather than late-onset ischemic stroke.^4^

We employed the two-sample MR method to assess the causal effects of migraine and its subtypes on early-onset ischemic stroke (Figure 1).

**Figure 1.**
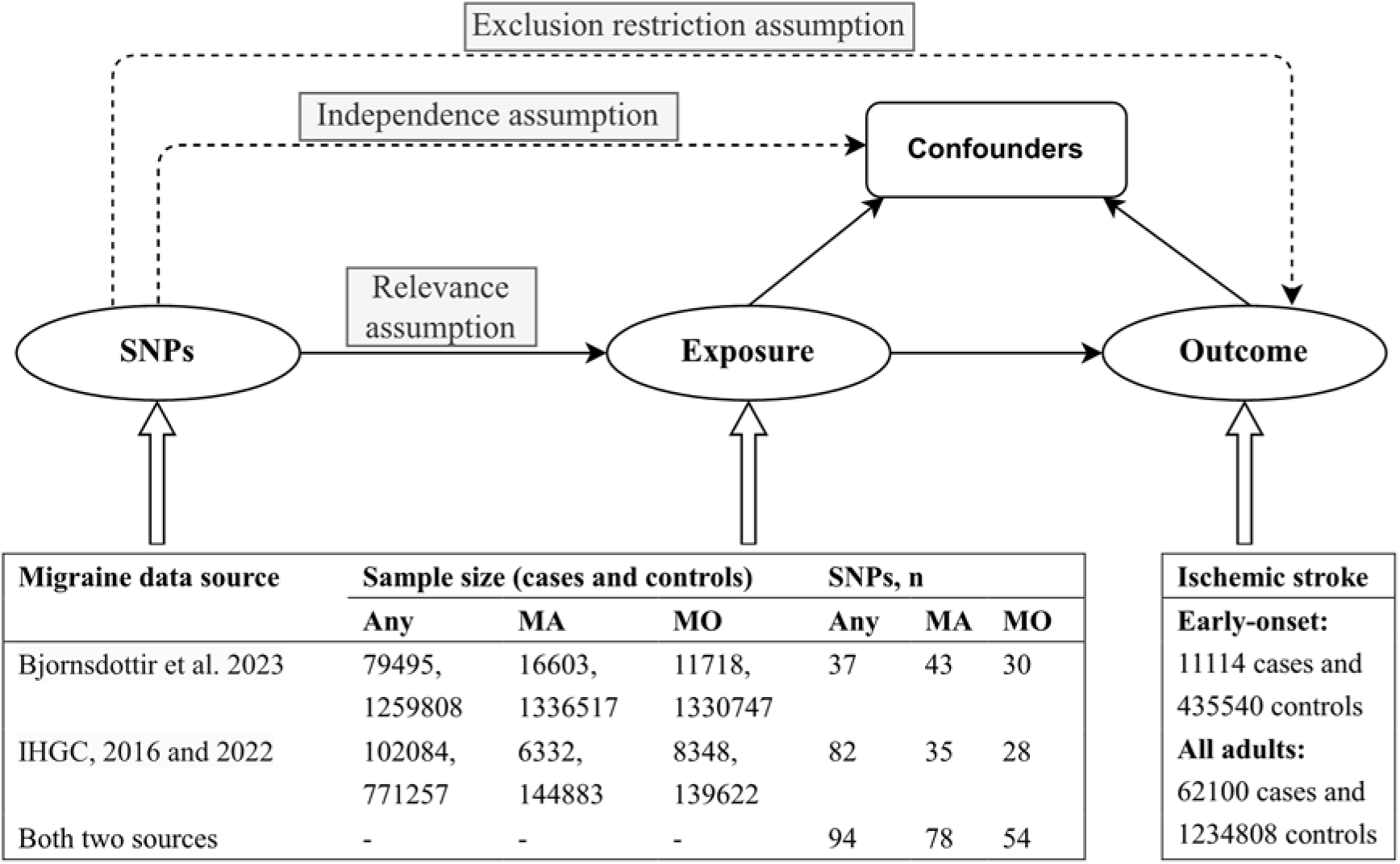
Design of the present study. In MR study, three key assumptions are crucial: the relevance assumption, stating that the genetic variant strongly associates with the exposure; the independence assumption, ensuring the genetic variant’s independence from confounders; and the exclusion restriction assumption, which dictates that the genetic variant solely affects the outcome through the exposure factor. For instrumental SNPs used in both data sources, we first combined un-clumped SNPs from the two sources and then conducted the clumping procedure. **Abbreviations**: Any, any migraine; MA, migraine with aura; MO, migraine without aura; SNP, single nucleotide polymorphism; IHGC, International Headache Genetics Consortium.

## Methods

### Summary data for migraine

Three sets of instrumental SNPs were used. The first set of instrumental SNPs were chosen from the genome-wide association study (GWAS) performed by Bjornsdottir et al recently (Supplementary Table 1).^5^ The second set of instrumental SNPs were chosen from two GWASs performed by the International Headache Genetics Consortium (IHGC) (Supplementary Table 2).^6, 7^ The third set of instrumental SNPs were obtained by combining the SNPs from both the two sources together to carry out the clumping procedure. Significant variants at the genome-wide level (p-value < 5 × 10^-8^) were utilized as instrumental variables for any migraine. As there is a limited number of variants with a p-value < 5 × 10^-8^ for migraine subtypes, similar to the previous MR studies associated with migraine subtypes, the threshold for instrumental variables was relaxed to < 1 × 10^-5^.^2, 3, 8^ The clumping procedure was carried out by employing an r^2^ value of 0.01 and a physical window of 10 MB. Supplementary Figure 1 shows the flow diagram of instrumental variables selection. SNPs associated with cardiometabolic risk factors were excluded (Supplementary Table 3 and Supplementary Table 4).^9^

### Summary data for early-onset ischemic stroke

Jaworek et al conducted a GWAS meta-analysis on early-onset ischemic stroke, with individuals ranging in age from 18 to 59 years.^10^ We employed summary data specific to Europeans, which encompassed 11,114 cases and 435,540 controls, to carry out the MR analysis. To make a comparison, we also carried out MR analyses by utilizing the summary data of ischemic stroke which is not specifically for early onset.^11^ If instrumental SNPs are not found in the outcome data, proxy SNPs (with an r^2^ > 0.8) will be used if available.

### Mendelian Randomization Analysis

We employed the random-effects inverse variance weighted method as the primary analysis approach. We also utilized MR-Egger, weighted median, and mode-based MR methods to account for the impact of horizontal pleiotropy in MR studies. Additionally, we employed the MR Pleiotropy RESidual Sum and Outlier approach to identify potential outliers.

We carried out MR analysis by utilizing the TwoSampleMR R package within R (version 3.6.1).^12^ We estimated the genetic correlation between migraine and ischemic stroke using the LD score software (LDSC v1.0.1).^13^

## Results

Supplementary Table 5 show the characteristics of SNP included. The MR analysis demonstrated that overall migraine and migraine without aura were not related to the risk of early-onset ischemic stroke (Figure 2 and Supplementary Table 6).

**Figure 2.**
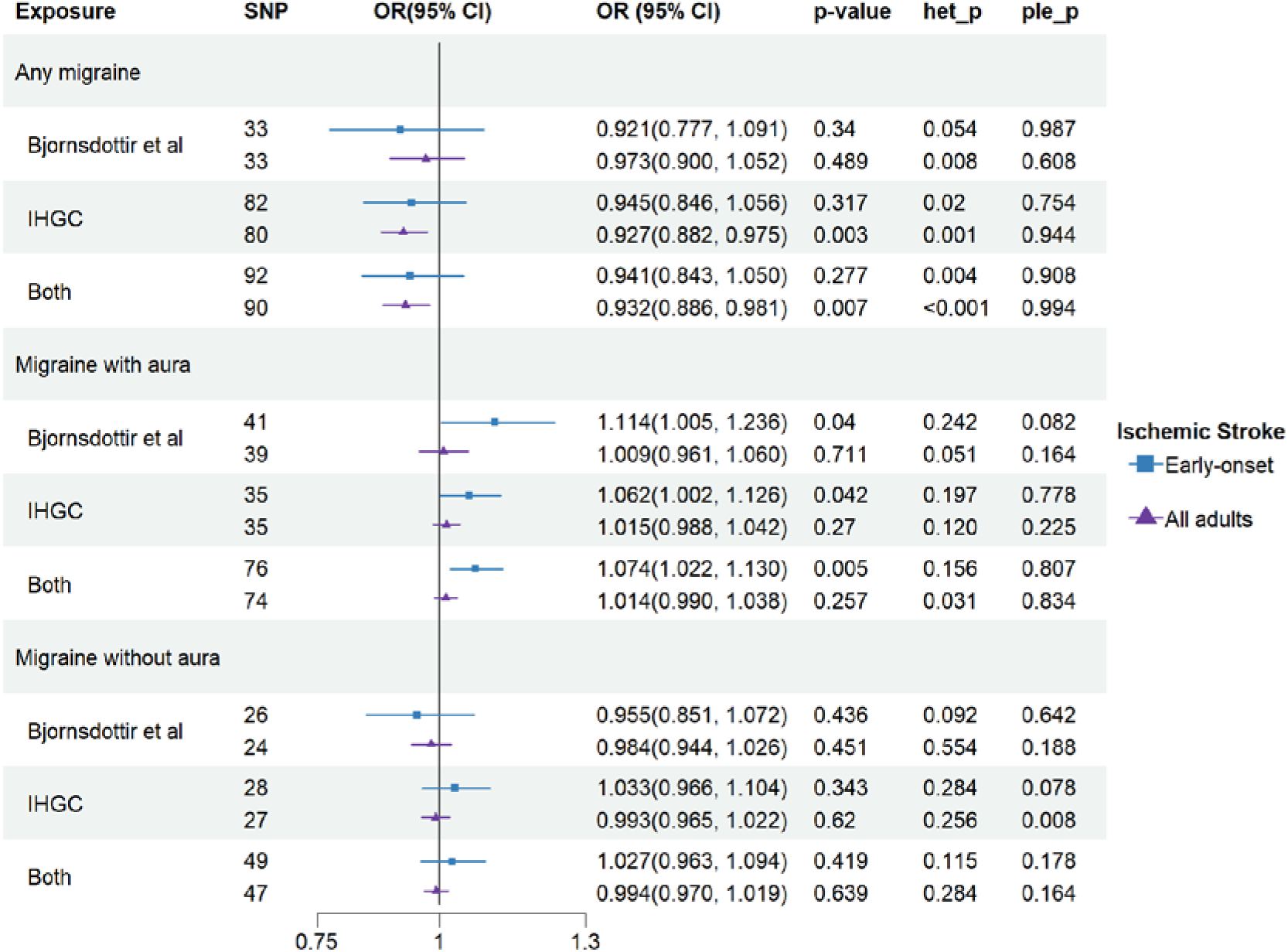
Inverse variance weighted estimates of migraine on risk of ischemic stroke. Het_p indicates p-value of heterogeneity test. Ple_p indicates p-value of pleiotropy test. **Abbreviations**: SNP, single nucleotide polymorphism; IHGC, International Headache Genetics Consortium; OR, odds ratio; CI, confidence interval.

For migraine with aura, the odds ratios (OR) based on instrumental SNPs from Bjornsdottir et al., the International Headache Genetics Consortium, and both sources were 1.114 (95% confidence interval [CI] = 1.005 to 1.236, p-value = 0.040), 1.062 (95% CI = 1.002 to 1.126, p-value = 0.042), and 1.074 (95% CI = 1.022 to 1.130, p-value = 0.005), respectively (Figure 2, Supplementary Figure 2, Supplementary Figure 3, and Supplementary Table 6). There was no evidence of heterogeneity or horizontal pleiotropy. The MR Pleiotropy RESidual Sum and Outlier method did not detect any outliers. Leave-one-out analysis demonstrated that the results were not driven by any single SNP (Supplementary Figure 4). The Steiger test did not find reverse causal SNPs. Sensitivity analyses showed similar results (Supplementary Figure 5, Supplementary Table 7, and Supplementary Table 8). Lastly, migraine with aura was not related to the risk of all adult ischemic strokes (Figure 2).

Due to the limited sample size, MR analysis did not find consistent associations for early-onset ischemic stroke subtypes (Supplementary Figure 6). Reverse MR analysis showed that early-onset ischemic stroke was not associated with the risk of any migraine and its subtypes (Supplementary Figure 7).

Linkage disequilibrium score regression analysis showed a positive genetic correlation between migraine with aura and early-onset ischemic stroke (rg = 0.208, se = 0.086, p-value = 0.016) and a negative genetic correlation between migraine without aura and early-onset ischemic stroke (rg = -0.197, se = 0.099, p-value = 0.047). The genetic correlation between migraine with aura and ischemic stroke of all adults attenuated to 0.154 (se = 0.064, p-value = 0.015).

## Discussion

Previous studies suggested that migraine, especially migraine with aura, might be associated with ischemic stroke in young adults, but not in older.^4^ The age-related differential risk might be explained by the fact that, migraine and age-related risk factors such as hypertension might have shared underlying mechanisms. In older individuals, age-related risk factors condition may saturate the biological mechanisms that ultimately lead to ischemic stroke, so that there are few open pathways for migraine to affect ischemic stroke.^14^

There were several possible underlying mechanisms for the association between migraine and ischemic stroke. First, previous studies have revealed that migraine shares genetic basis with cardiometabolic risk factors and thrombosis.^10^ On the other hand, migraine with aura shows a higher prevalence of patent foramen ovale compared to migraine without aura, potentially elevating the risk of paradoxical embolism. Third, spreading depolarizations likely play a pivotal role in the increased risk of ischemic stroke in individuals with migraine, especially those with migraine accompanied by aura. Last, additional pathophysiological mechanisms might involve inflammatory markers, endothelial and vascular indicators, as well as sex hormones.^15^

The present study has several limitations. First, due to the unavailability of sex-specific summary data, we were unable to carry out sex-stratified analysis. Second, since only a few SNPs associated with migraine with aura reach genome-wide significance, we employed a more lenient threshold for instrumental variables. Third, due to the limited sample size of stroke subtypes, MR analysis did not identify consistent associations among the three migraine data sources. Lastly, the conclusions drawn from this study rely on summary data from European populations, potentially limiting their generalizability to other ethnic groups.

## Conclusion

In conclusion, this study confirmed the causal effect of migraine with aura on early-onset ischemic stroke risk. Further studies are needed to confirm the findings and to address the limitations of this study.

## Availability of data

The supporting data for this study can be found in the supplementary files of the article as well as in the referenced studies within this article.

Summary data for migraine by Bjornsdottir et al. were obtained from https://www.decode.com/summarydata/. Summary data for migraine by International Headache Genetics Consortium were obtained from https://www.headachegenetics.org/datasets-cohorts. Summary data for early-onset ischemic stroke were obtained from the Cerebrovascular Disease Knowledge Portal (https://cd.hugeamp.org/).

## Data Availability

The supporting data for this study can be found in the supplementary files of the article as well as in the referenced studies within this article.
Summary data for migraine by Bjornsdottir et al. were obtained from https://www.decode.com/summarydata/. Summary data for migraine by International Headache Genetics Consortium were obtained from https://www.headachegenetics.org/datasets-cohorts. Summary data for early-onset ischemic stroke were obtained from the Cerebrovascular Disease Knowledge Portal (https://cd.hugeamp.org/).

https://www.headachegenetics.org/datasets-cohorts

https://cd.hugeamp.org/

## Acknowledgments

We would like to acknowledge the participants and investigators of the included genome wide association studies.

---

**Supplementary Figure 1.**
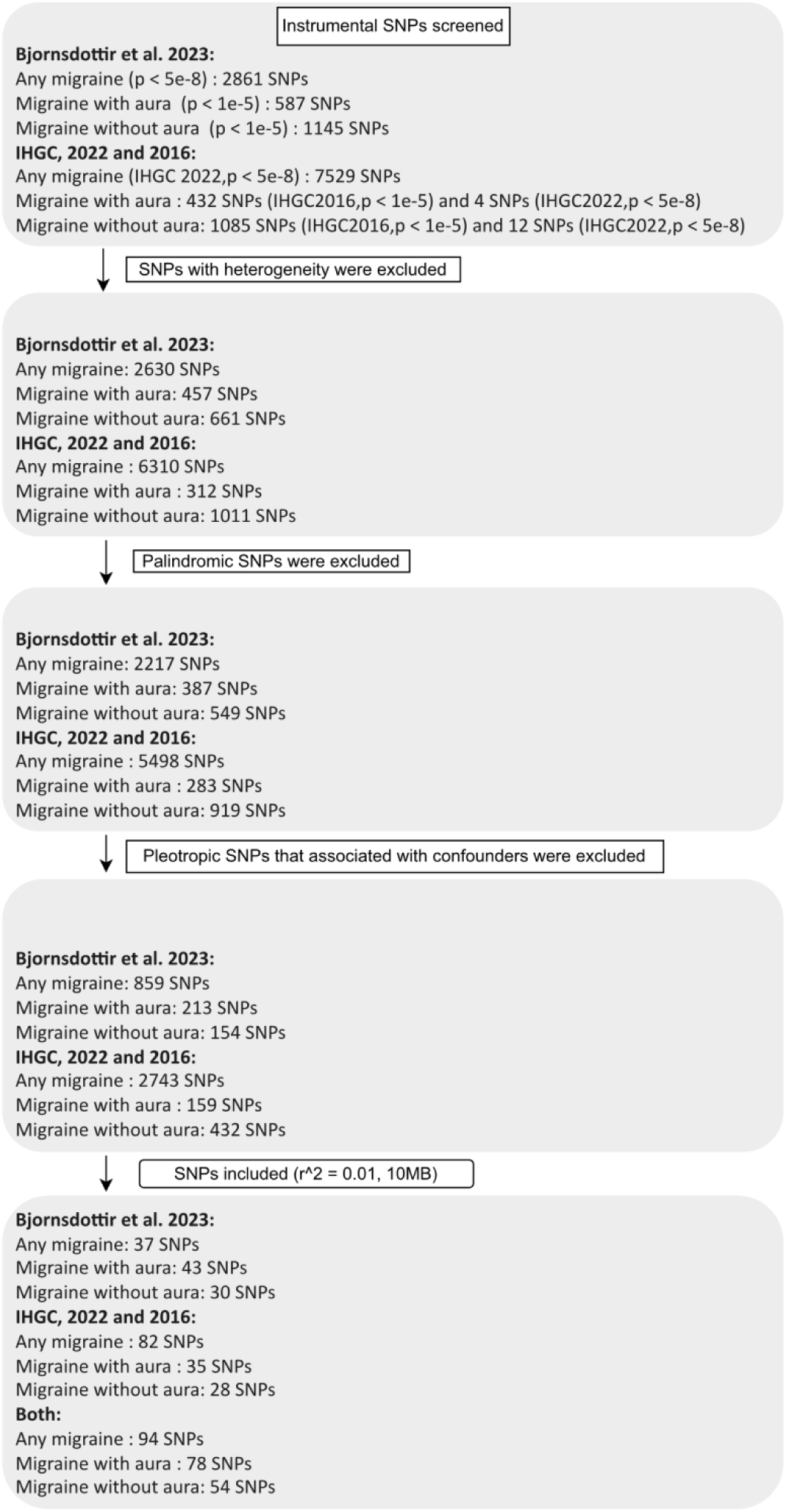
Flow diagram of instrumental variables selection. Significant single nucleotide polymorphisms (SNPs) at the genome-wide level (with a p-value < 5 × 10^-8^) were utilized as instrumental variables for any migraine. However, due to a limited number of SNPs with a p-value < 5 × 10^-8^ for migraine with aura and migraine without aura, the threshold for instrumental variables was relaxed to < 1 × 10^-5^.^1, 2^ SNPs that are absent in the 1000G v3 reference panel of the European population were excluded before being screened. Heterogeneity was determined to exist when the I^2^ exceeded 50% or the p-value of the Q statistic was less than 0.05. Palindromic SNPs were excluded since the effect allele frequency was missing in the outcome data. For the instrumental SNPs from both two sources, they were obtained through clumping on all the SNPs detailed in the box which is next to the last one. All instrumental SNPs exhibit an F statistics value greater than 10, and this was arrived at by computing beta2 divided by se2. References 1. Lee KJ, Lee SJ, Bae HJ, Sung J. Exploring the causal inference of migraine on stroke: A Mendelian randomization study. *Eur J Neurol* 2022;29(1):335-338. doi: 10.1111/ene.15101 2. Wu XP, Niu PP, Liu H. Association between migraine and venous thromboembolism: a Mendelian randomization and genetic correlation study. *Front Genet* 2024;15:1272599. doi: 10.3389/fgene.2024.1272599

**Supplementary Figure 2.**
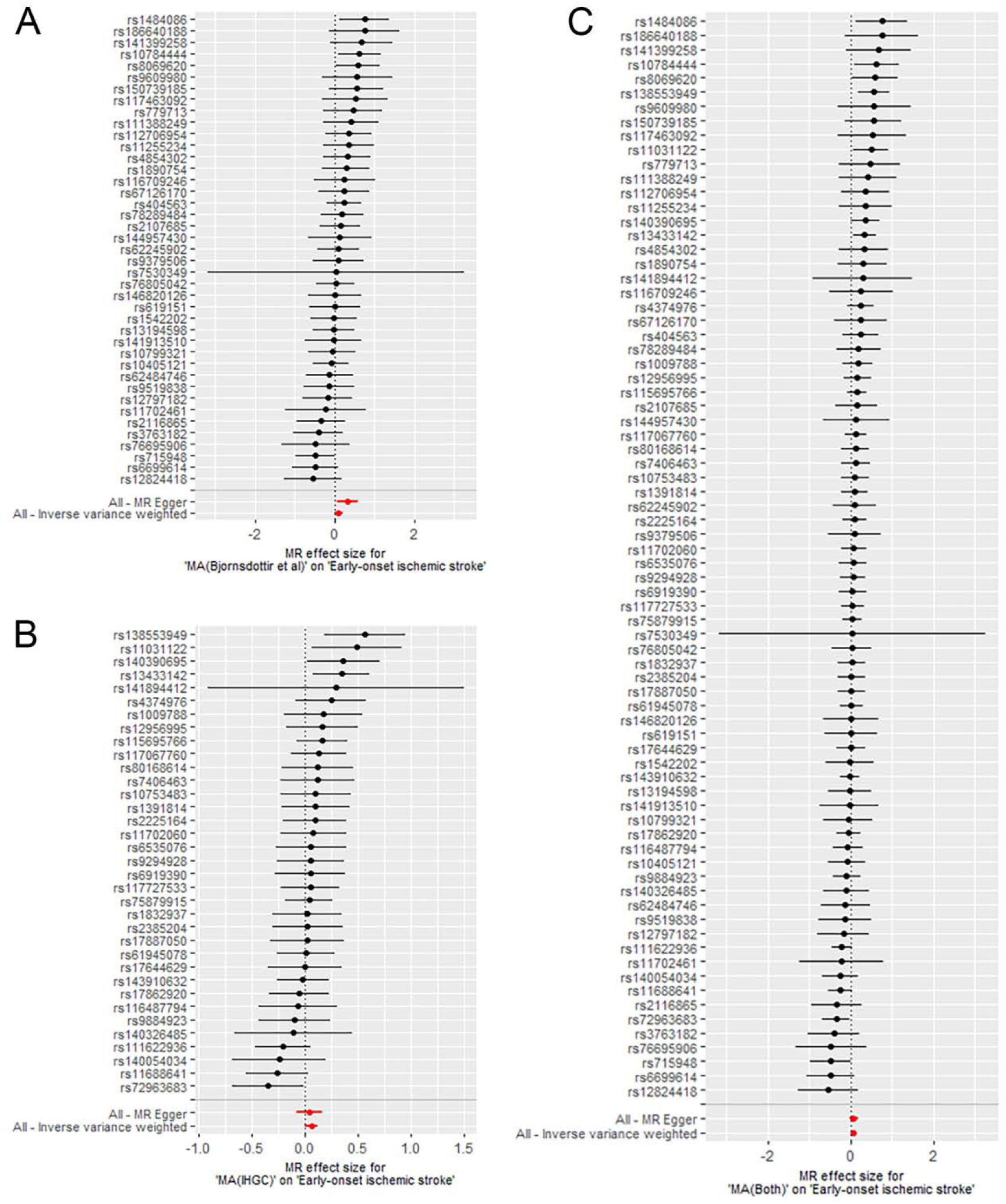
Forest plots of the effect of migraine with aura on early-onset ischemic stroke. The horizontal axis represents the estimate of migraine with aura on early-onset ischemic stroke. A: instrumental SNPs based on study by Bjornsdottir et al. 2023 B: instrumental SNPs based on study by IHGC C: instrumental SNPs based on both two sources.

**Supplementary Figure 3.**
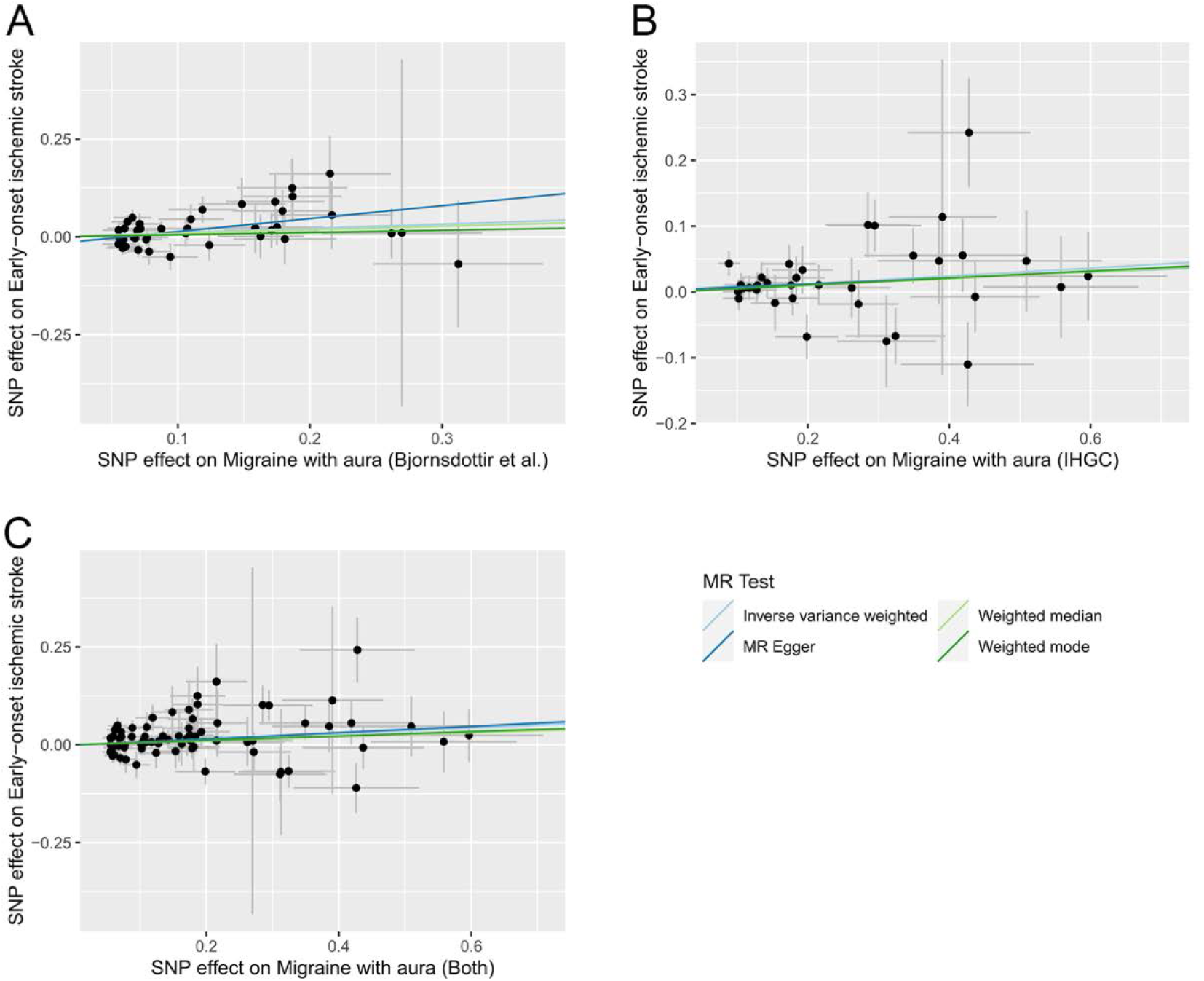
Scatter plots of SNP effect on migraine with aura and SNP effect on early-onset ischemic stroke. A: instrumental SNPs based on study by Bjornsdottir et al. 2023; B: instrumental SNPs based on study by IHGC; C: instrumental SNPs based on both two sources. Abbreviations: SNP, single nucleotide polymorphism; IHGC, International Headache Genetics Consortium.

**Supplementary Figure 4.**
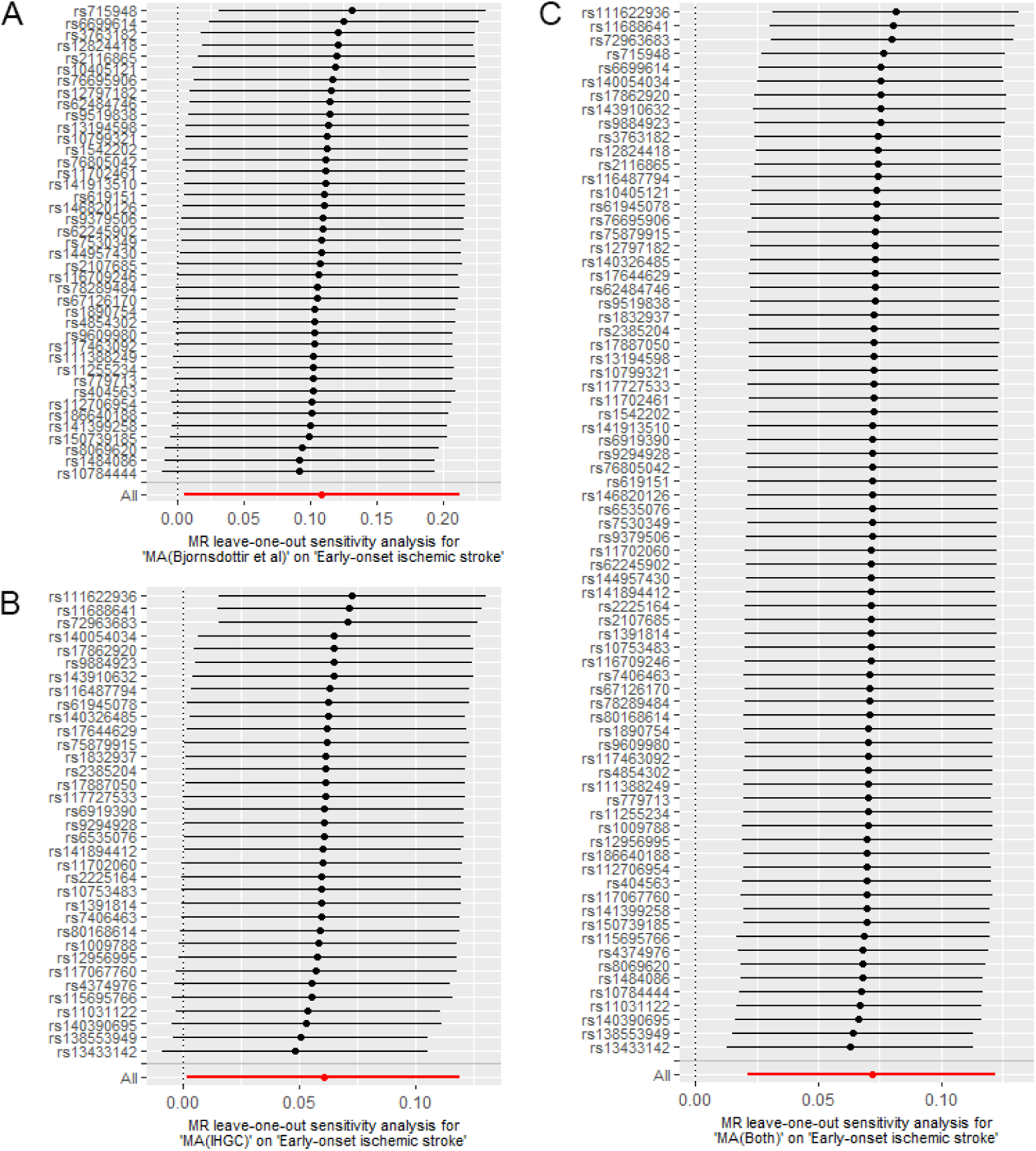
Leave-one-out sensitivity analysis of the effect of migraine with aura on early-onset ischemic stroke. A: instrumental SNPs based on study by Bjornsdottir et al. 2023 B: instrumental SNPs based on study by IHGC C: instrumental SNPs based on both two sources.

**Supplementary Figure 5.**
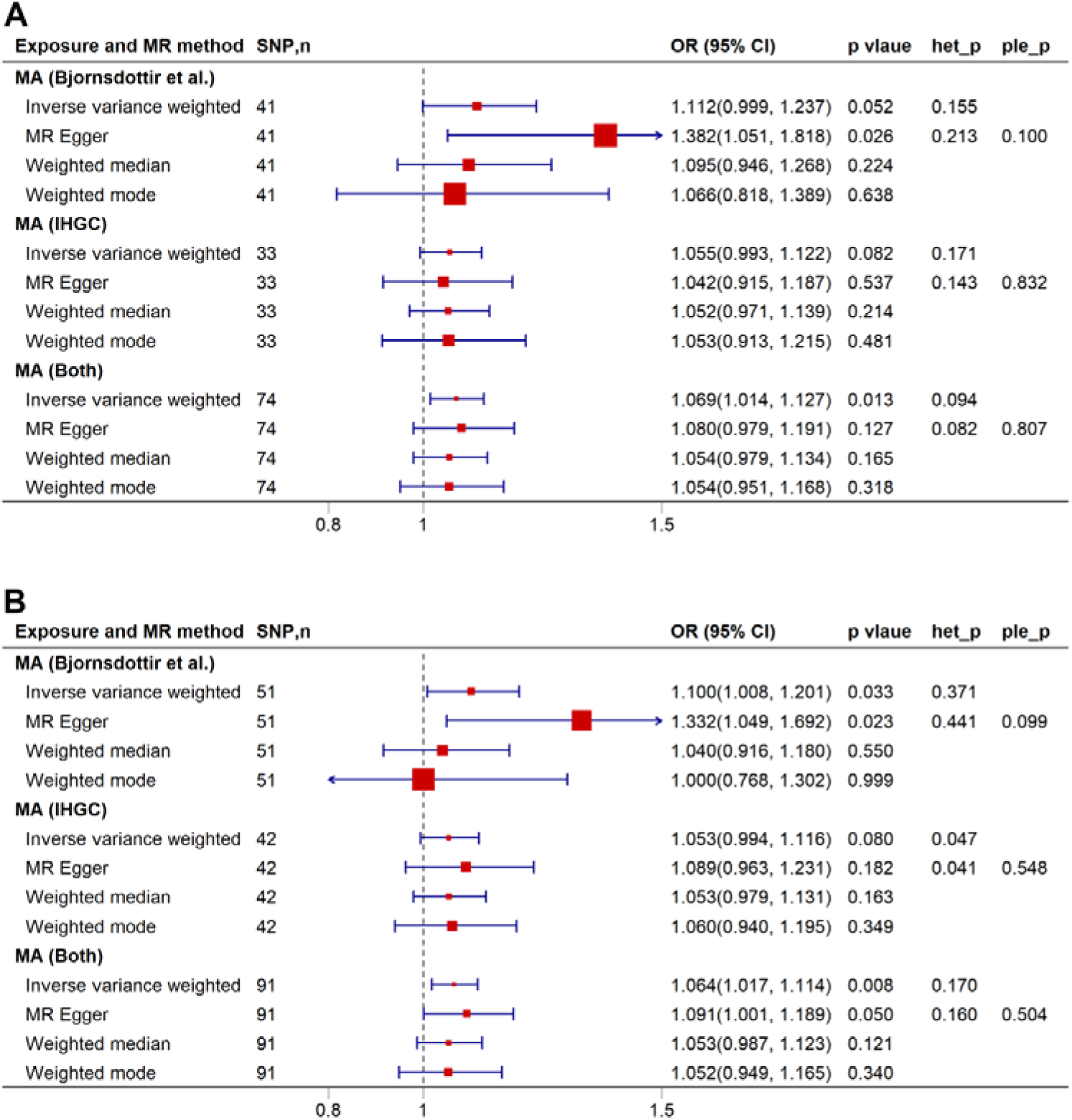
Sensitivity analyses of migraine with aura on early-onset ischemic stroke. A: Sensitivity analysis was conducted by exclusively relying on SNPs that exhibited low evidence of heterogeneity across different stroke cohorts (with I^2^ less than 50% and a p-value of the Q statistic greater than 0.05) B: Sensitivity analysis was conducted using SNPs, without consideration of whether they were associated with potential confounders. Het_p indicates p-value of heterogeneity test. Ple_p indicates p-value of pleiotropy test. Abbreviations: SNP, single nucleotide polymorphism; IHGC, International Headache Genetics Consortium; OR, odds ratio; CI, confidence interval; MA, migraine with aura.

**Supplementary Figure 6.**
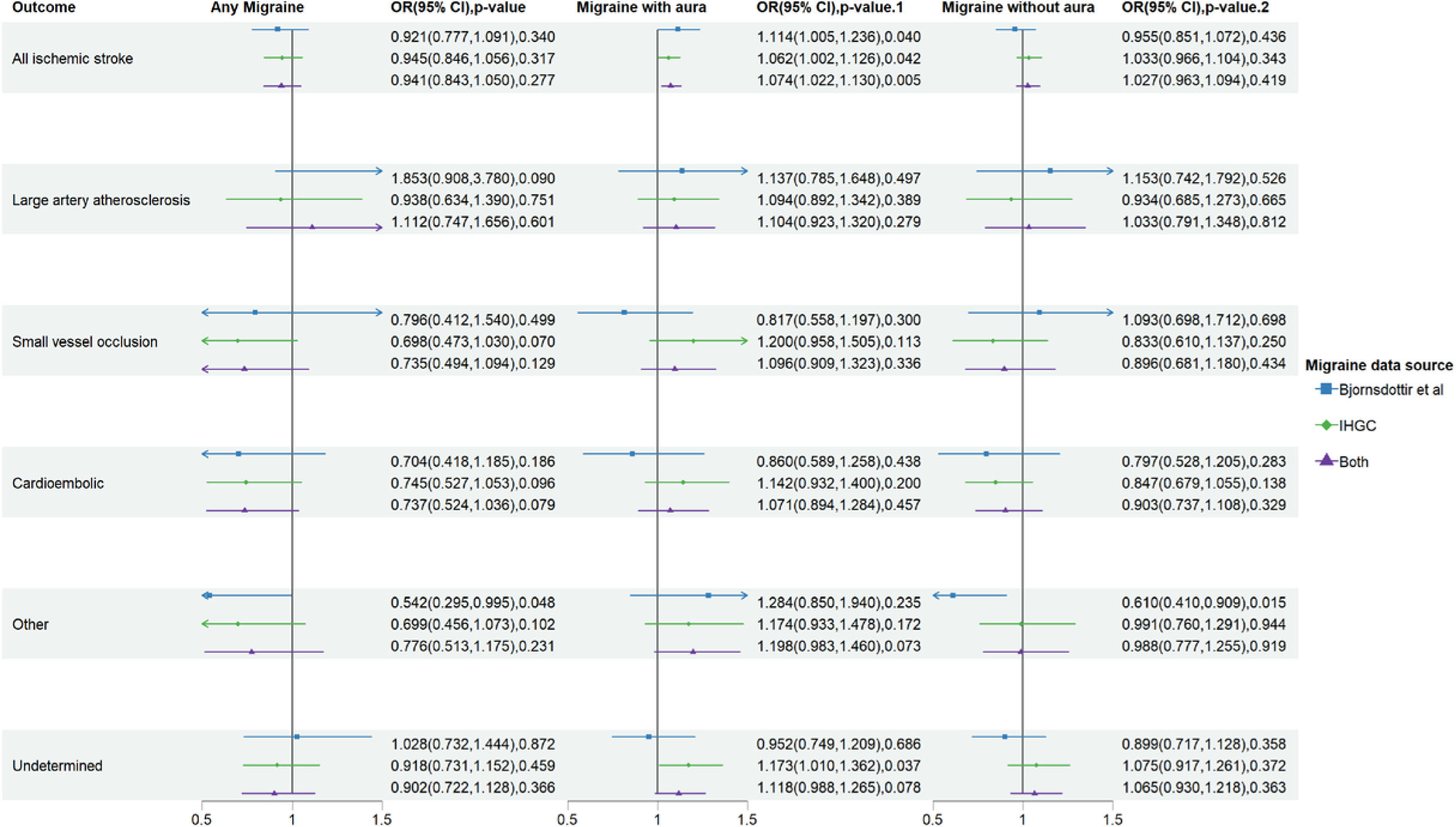
Inverse variance weighted estimates of migraine on risk of early-onset ischemic stroke subtypes. Abbreviations: MR, IHGC, International Headache Genetics Consortium; OR, odds ratio; CI, confidence interval.

**Supplementary Figure 7.**
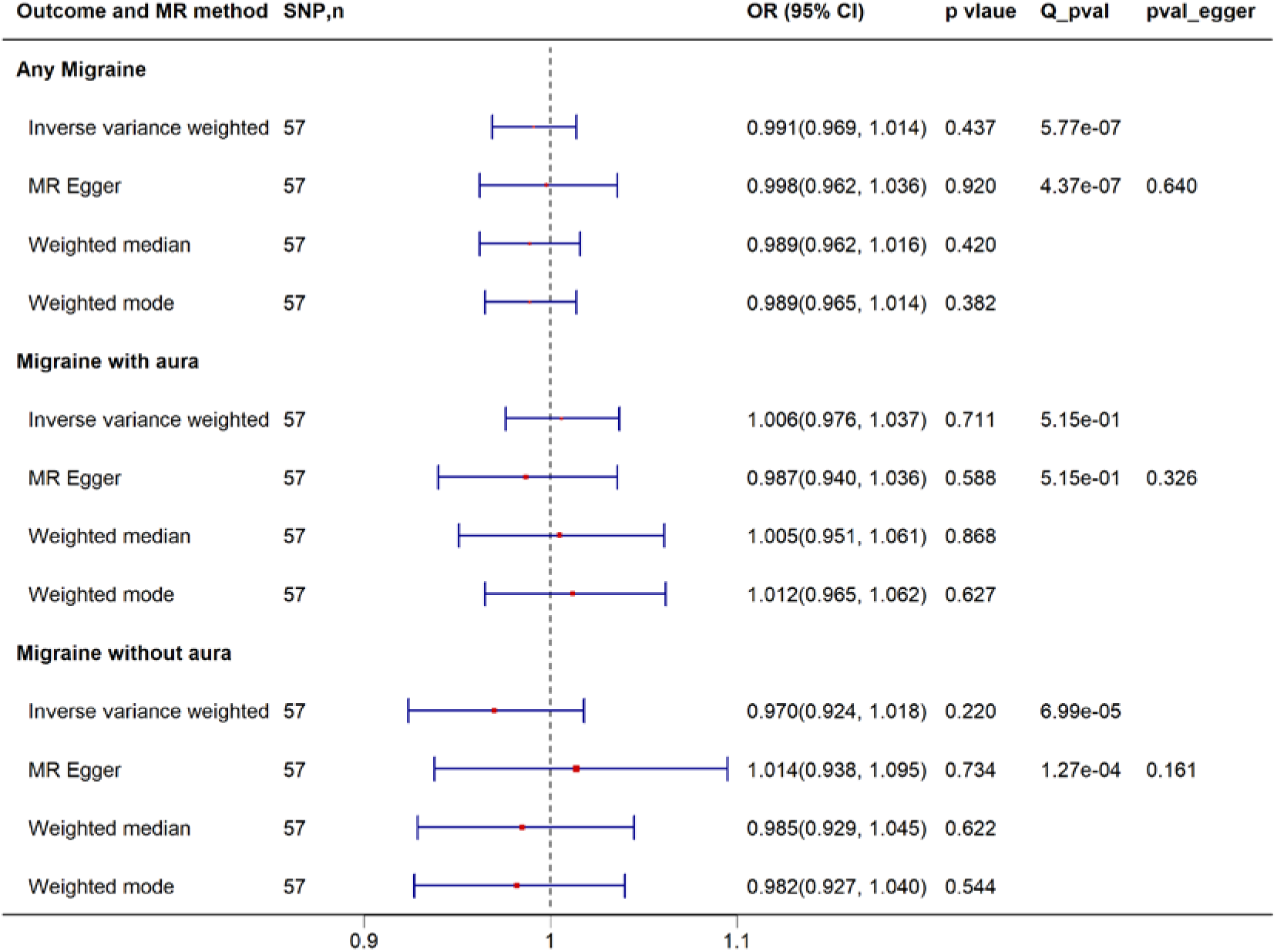
Reverse Mendelian randomization analysis of early-onset ischemic stroke on risk of migraine. Due to a limited number of SNPs with a p-value < 5 × 10^-8^ for early-onset ischemic stroke, the threshold for instrumental variables was relaxed to < 1 × 10^-5^ Abbreviations: MR, Mendelian randomization; SNP, single nucleotide polymorphism; OR, odds ratio; CI, confidence interval.

